# *Mycobacterium tuberculosis*-specific cytokine responses of infants born to mothers with active tuberculosis in Uganda

**DOI:** 10.1101/2024.10.23.24315978

**Authors:** Diana Sitenda, Phillip Ssekamatte, Rose Nakavuma, Andrew Peter Kyazze, Felix Bongomin, Joseph Baluku, Rose Nabatanzi, Davis Kibirige, Stephen Cose, Irene Andia-Biraro, Annettee Nakimuli

**Affiliations:** Department of Immunology and Molecular Biology, School of Biomedical Sciences, College of Health Sciences, Makerere University, Kampala, Uganda; Tuberculosis and comorbidities research consortium, Kampala, Uganda.; Department of Medical Microbiology and Immunology, Gulu University (GU), Gulu, Uganda; Department of Medicine, Uganda Martyrs’ Hospital Lubaga, Kampala, Uganda; Medical Research Council/ Uganda Virus Research Institute and London School of Hygiene and Tropical Medicine, Entebbe, Uganda; Department of Internal Medicine, School of Medicine, College of Health Sciences, Makerere University, Kampala, Uganda; Department of Obstetrics and Gynaecology, School of Medicine, College of Health Sciences, Makerere University, Kampala, Uganda

**Keywords:** Cytokine responses, infants, active TB, pregnancy, Uganda

## Abstract

**Background:** Immunizing infants with various vaccines, including *Bacillus Calmette–Guérin* (BCG), Diphtheria-Pertusis-Tetanus (DPT), and measles, aims to enhance immunity. In instances where vaccine responses have been reported to be compromised, individuals are prone to infection. The BCG vaccine, for example, induces strong type 1 immune responses, particularly interferon-gamma (IFN-γ) expression, that are essential for protection against *Mycobacterium tuberculosis* (*Mtb)*. However, there is scanty evidence on whether this effect is established or sustained when infants are exposed to *Mtb* either in utero or after birth. We compared TB-specific cytokine responses for IFN-γ, interleukin (IL)-2 (IL-2), tumour necrosis factor-alpha (TNF-α), IL-17A, and Granulocyte-macrophage colony-stimulating factor (GM-CSF) using supernatants harvested from QFT-Plus Blood Collection Tubes.

**Methods:** This cross-sectional study compared 22 infants born to mothers with bacteriologically confirmed active tuberculosis (TB), defined as TB exposed or cases, to 20 infants born to mothers without active TB, defined as TB non-exposed or controls. Plasma harvested from the QFT-plus tubes (TB1 and TB2) was used to perform a 5-plex Luminex assay using the LX 100/200 Luminex machine and measured in pg/mL. Data was analysed using R (v.4.4.1). The Mann-Whitney U test was used to determine statistical significance at a p-value less than 0.05 and a 95% confidence interval. Data was expressed as median and interquartile ranges (IQR).

**Results:** TB-exposed infants showed IFNγ responses were slightly higher among TB-exposed infants compared to non-exposed (Medians (IQR): 15.49 (14.58-16.49) versus 14.96 (14.60-16.60), p=0.68, respectively. There was a strong expression of total IL-17A among TB-exposed compared to non-exposed 11.91 (10.89-13.50) versus 10.69 (10.17-11.64), p=0.035. We observed no differences in IL-2, TNF, and GM-CSF responses.

**Conclusion:** TB exposure among infants slightly alters their *Mtb*-specific cytokine responses, especially IL-17A cytokine responses. This suggests possible ongoing *Mtb* infection among TB-exposed infants. Follow-up studies of such infants are necessary to assess their risk of future TB infection and disease and the potential need for TB chemoprophylaxis.

## Introduction

Tuberculosis (TB) remains one of the top ten causes of death and the leading cause of death from a single infectious agent on a global scale [1]. One-third of the population is believed to be infected with *Mycobacterium tuberculosis (Mtb)*, with the biggest percentage of the population developing a latent TB infection and about 10% developing active TB during their lifetime [2]. However, the risk of developing active TB is higher among immunocompromised individuals such as young children (aged 5 years and below), HIV infected, and the elderly [3], [4] & [5]. According to Verkuijl *et*., *al* 2023, over 1.25 million children and young adolescents were infected with TB in 2022, of whom 47% were children below 5 years of age [6]. In addition, the age group below 5 years alone is reported to contribute about 12% of the total TB incidence, at 10.6 million [6]. Congenital TB is rare; however, when it occurs, infants tend to have poor outcomes [7]. Mother-to-child transmission of TB may occur *in utero* through hematogenous spread via the umbilical vein and aspiration or swallowing of infected amniotic fluid [8]. TB exposure in utero among infants has been associated with outcomes including respiratory distress and very low birth weights [9].

IFN-γ is the key cytokine for a protective immune response against *Mtb* [10] and is produced by CD4^+^, CD8^+^ T cells, and NK cells. The IFN-γ augments antigen presentation, leading to the recruitment of CD4^+^ T-cells and/or cytotoxic CD8^+^ T-cells, which participate in mycobacterial killing and prevent exhaustion of memory T cells [11]. Other cytokines associated with the T helper (Th) 1 and Th17 response, such as TNF-α, IL-2, and IL-17, also play an essential role in protection from TB [12]. Similarly, IL-17 acts as an effector molecule after *Bacille Calmette-Guérin* (BCG) vaccination and *Mtb* infection to protect humans against TB. IL-17 fights *Mtb* by inducing inflammation and chemokines [12]. In addition, IL-17 enhances the accumulation of polymorphic and mononuclear cells, which could kill TB [13]. GM-CSF is, on the other hand, expressed within human TB granulomas, enhancing the recruitment of granulocytes and macrophages [14].

The World Health Organisation recommends that infants are vaccinated with vaccines such as *Bacillus Calmette–Guérin* (BCG) to fight early-life infections [15] and in Uganda today, the extended program on immunization recommends over 11 vaccines during childhood, including BCG, Diphtheria-Pertusis-Tetanus (DPT), and measles. BCG-induced protection against TB is implicated by Th1 cells through its production of IFN gamma [16].

This study aimed to explore the hypothesis that infants born to mothers with active TB will exhibit heightened TB-specific Th1 cytokine responses, particularly IL-17, IFN-γ, TNF-α, IL-2, and GM-CSF responses, compared to those born to mothers without active TB. **Methods**

### Description of study setting and participants

This cross-sectional study carried out in October 2021 enrolled 35 women with bacteriologically confirmed active TB as cases and 33 women without TB as controls matched for maternal and gestational age from antenatal (ANC) and postnatal (PNC) clinics at three major health facilities in Kampala, Uganda: Kasangati Health Center IV, Kisenyi Health Center IV, and Kawempe National Referral Hospital. We, however, considered 22 infants born to mothers who had bacteriologically confirmed active TB (TB-exposed), also termed cases, and 20 infants born to mothers without TB (TB non-exposed), also termed controls. These infants were matched for age and gestation. Upon delivery, the infants were assented and enrolled in the study.

### Study procedures

These infants donated 4ml of blood at baseline for latent TB infection testing using the QFT-plus assay. Each of the four QuantiFERON tubes; Nil, TB1, TB2, and Mitogen, were aseptically filled with 1mL of blood, inverted up and down to mix the blood with stimulants of ESAT-6 and CFP-10 coated on the walls of the TB1 and TB2 tubes, and the Phytohemagglutinin coated on the walls of the mitogen tube. This inversion was done ten times, and the samples were immediately incubated at 37ºC in an incubator and timed to complete incubation following 16 to 24 hours. The sample tubes were removed from the incubator and spun at 4000 RPM for 10 minutes. The supernatants were harvested and stored at -80□ in a freezer at the Immunology Laboratory Makerere University College of Health Sciences (MakCHS).

### Luminex assay

Infants QFT supernatant samples from TB1 and TB2 of the QFT tubes were retrieved and transferred to the fridge at 4□ overnight. The samples and the two 5-plex Luminex kits were thawed for 1 hour at room temperature. Protocols were designed in the xPONENT program for the LX 100/200 Luminex machine under the guide of the kits’ certificates of analysis, ensuring bead regions for the different cytokines were selected. The assay was performed according to the manufacturer’s instructions for the commercial kit: R&D systems-biotechne; Human Magnetic Luminex Assays.

50μl undiluted sample, standards, and Calibrator diluent (for the blank wells) were loaded in duplicate into the selected wells. This was followed by the addition of 50μl of reconstituted microparticle cocktail to all wells, using a multichannel pipette. Plates were incubated at room temperature, for 2 hours, on a horizontal orbital plate shaker at 800rpm. The plates were then manually washed 3 times with 100μl of 1X wash buffer using a Luminex magnetic plate. This was followed by the addition of 50μl of reconstituted biotin-antibody cocktail to all wells, incubated for 1 hour at room temperature, on a horizontal orbital plate shaker at 800rpm. The plates were washed again, and 50μl of diluted streptavidin-PE were added to all wells and the incubation procedure was repeated for 30 minutes. The plates were washed and finally, 100 μl of wash buffer were added to all wells and incubated for 2 minutes at room temperature on a shaker at 800rpm. The plates were then placed in the Milliplex Analyser (LX 100/200) to determine the bead count, mean fluorescence intensity, and concentrations of the *Mtb*-specific cytokines in the samples, using the Luminex xPONENT program version 4.3.309.1.

### Data analysis

Data was analysed using R (version 4.4.1) with the Mann-Whitney U test for statistical comparisons. Data was cleaned to remove missing values and outliers. The data was log_2_ transformed, and correlation coefficients were generated using Spearman pairwise correlation for correlation analysis. The data were depicted using a dot plot to illustrate single-sample distribution, a violin plot to characterise group distribution behaviour, and a box plot to present the confidence interval surrounding the median. Additionally, the box boundary indicates the interquartile range (IQR), while the whiskers represent 1.5 times the IQR. A P-value <0.05 was considered statistically significant at a 95% confidence level. Data were expressed as medians and interquartile ranges. The R package, ‘ggplot2’, was used to visualise the results for all plots.

## Results

### Participant demographic characteristics

Table 1 shows that TB-exposed infants (cases) had low haemoglobin concentrations(g/dL) compared to non-exposed infants (controls). The median and IQR for exposed versus non-exposed infants is 10.60 (9.58-10.90) versus 11.70 (11.40-13.10), p=0.046, respectively. In addition, the analysis revealed that TB-exposed infants had low birth weights compared to the non-exposed infants. The median and IQR for exposed versus non-exposed infants is 3.00 (2.00-3.17) versus 3.40 (2.70-4.20), p=0.035, respectively. Finally, other characteristics such as age and sex did not confound our findings. The p-values for age and sex are p=0.509 and 0.782, respectively, as shown in Table 1.

**Table 1:**
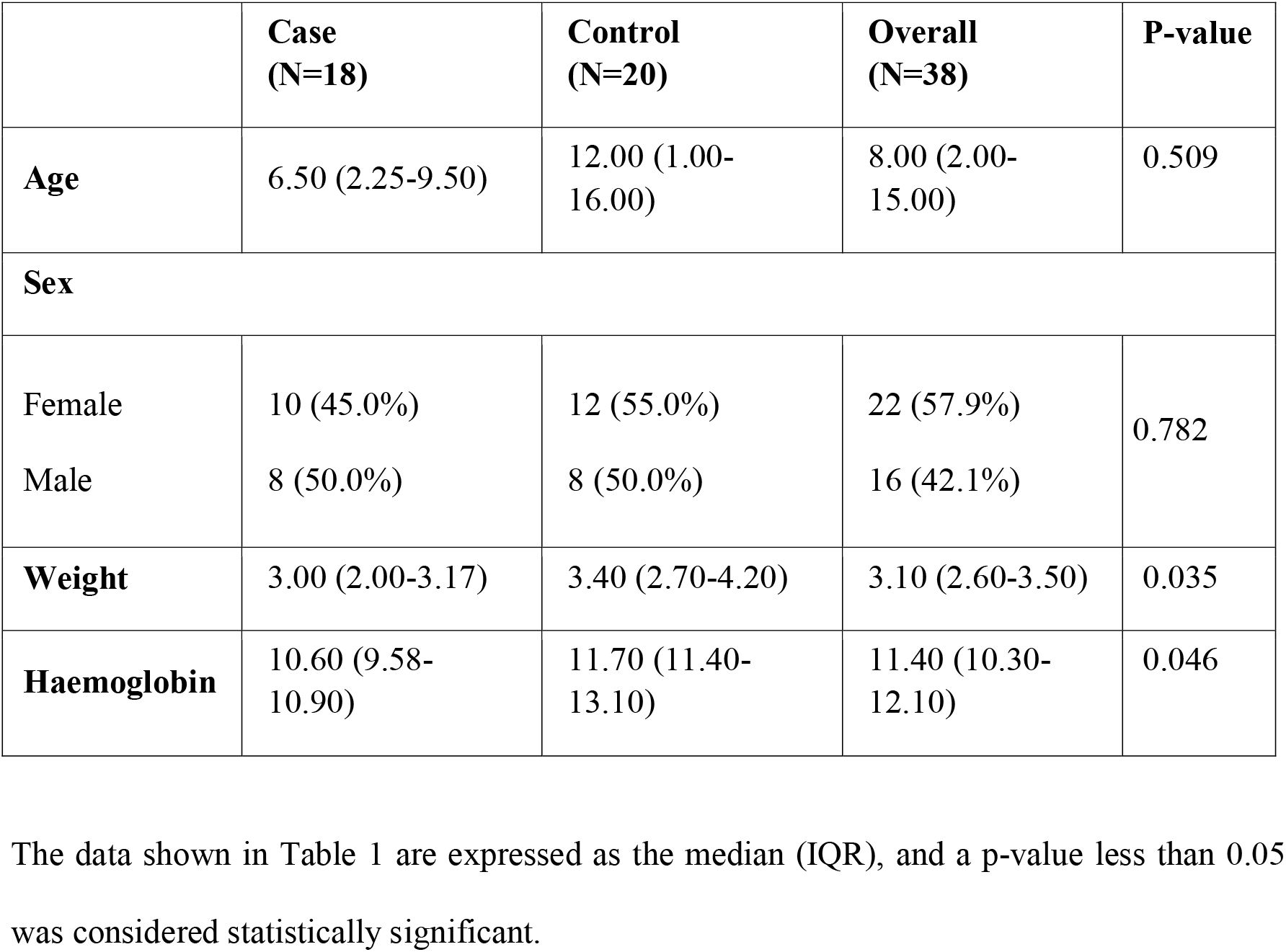
Characteristics of the study participants.

### TB-specific cytokine responses

#### Total cytokine responses (TB1 and TB2)

Cytokine responses were measured using the Luminex 5-plex assay for infants born to active TB mothers (TB-exposed) and infants born to mothers without TB (TB-non-exposed). We observed a significant increase in the total IL-17A cytokine response among TB-exposed infants versus TB-non-exposed infants: p=0.034, as shown in Figure 1A. The median and IQR are shown in Table 2.

**Table 2:** Cytokine response levels.

| Cytokines | TB-Exposed |  |  |  |  |  | TB Non-exposed |  |  |  |  |  |
| --- | --- | --- | --- | --- | --- | --- | --- | --- | --- | --- | --- | --- |
|  | Total<br>median<br>(IQR) | P-value | TB1<br>median<br>(IQR) | P-value | TB2<br>median<br>(IQR) | P-value | Total<br>median<br>(IQR) | P-value | TB1<br>median<br>(IQR) | P-value | TB2<br>median<br>(IQR) | P-value |
| IL-17A | 11.909<br>(10.888-13.504) | 0.035 | 5.575<br>(5.176-6.752) | 0.15 | 6.137<br>(5.282-6.752) | 0.049 | 10.690<br>(10.168-11.635) | 0.035 | 5.421<br>(4.928-5.956) | 0.15 | 5.326<br>(4.907-6.032) | 0.049 |
| GM-CSF | 14.042<br>(12.391-16.388) | 0.592 | 6.604<br>(5.766-8.096) | 0.195 | 7.505<br>(6.599-8.377) | 0.94 | 14.748<br>(13.099-16.659) | 0.592 | 7.342<br>(6.590-8.328) | 0.195 | 7.314<br>(6.574-8.572) | 0.94 |
| IFN-γ | 15.489<br>(14.577-16.488) | 0.677 | 7.903<br>(7.406-8.287) | 0.613 | 7.725<br>(7.430-8.302) | 0.623 | 14.962<br>(14.603-16.604) | 0.677 | 7.478<br>(7.166-8.302) | 0.613 | 7.505<br>(7.315-8.302) | 0.623 |
| TNF | 19.561<br>(18.684- | 0.753 | 9.477<br>(9.144- | 0.696 | 9.816<br>(9.477- | 0.489 | 19.439<br>(18.136- | 0.753 | 9.744<br>(8.882- | 0.696 | 9.650<br>(9.192- | 0.489 |
|  | 20.268) |  | 9.959) |  | 10.192<br>) |  | 20.339) |  | 10.051<br>) |  | 10.333<br>) |  |
| IL-2 | 13.412<br>(12.630-15.817) | 0.194 | 7.288<br>(6.216-7.908) | 0.175 | 6.437<br>(6.116-7.908) | 0.649 | 12.813<br>(12.075-13.897) | 0.194 | 6.371<br>(6.024-7.272) | 0.175 | 6.513<br>(6.053-7.281) | 0.649 |

Data was presented as median and IQR, a p-value less than 0.05 was considered statistically significant after performing the Man-Whitney U test. The total responses were determined by combining responses from both the TB1 and TB2 QFT tubes.

**Figure 1:**
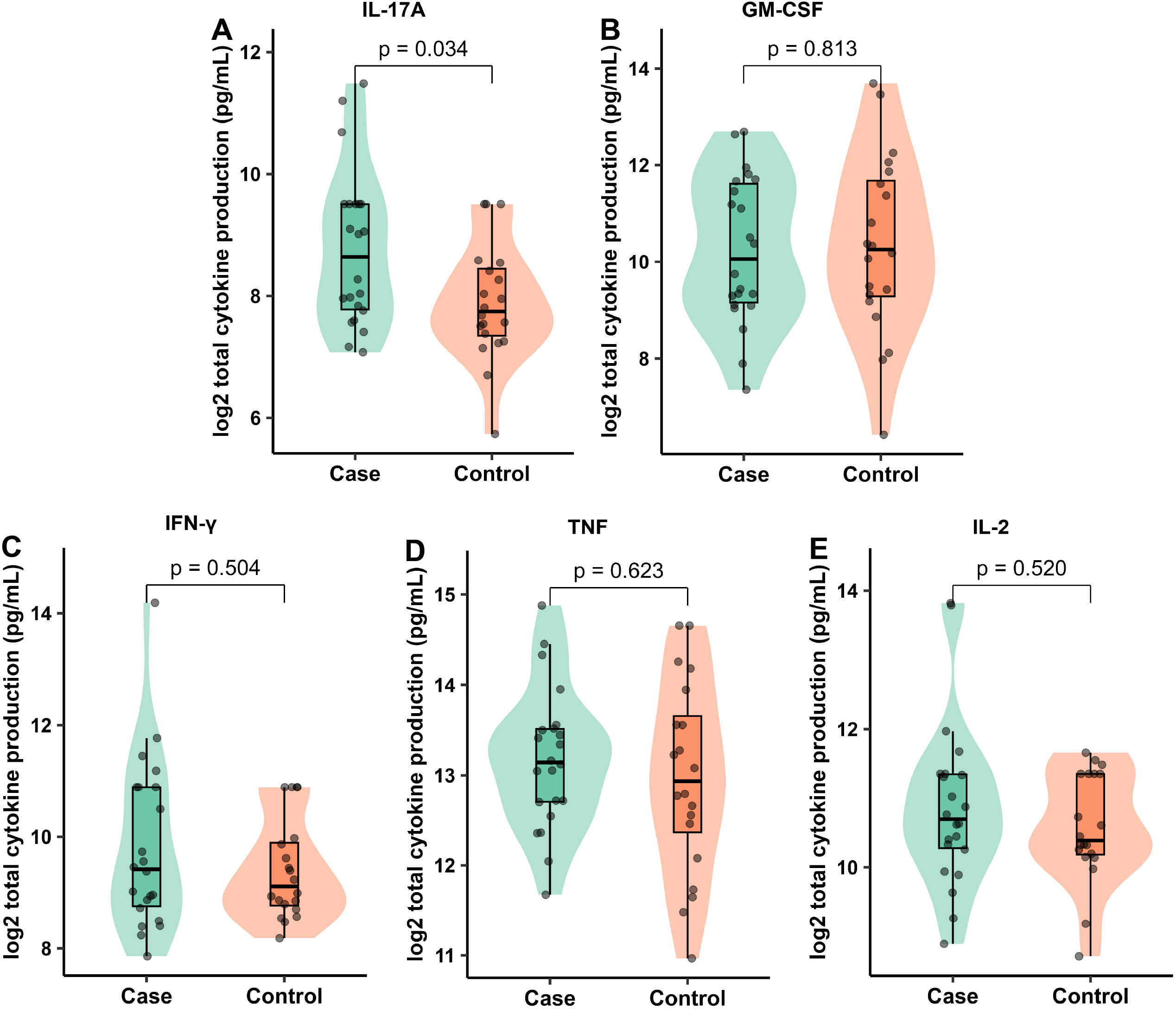
Total cytokine responses for IL-17A, GM-CSF, IFN γ, TNF-α, and IL-2 among TB-exposed and TB-non-exposed infants were measured by Luminex. Total responses for IL-17A(A), GM-CSF(B), IFN-γ(C), TNF-α(D), and IL-2(E). Box plots show data represented as median and interquartile ranges. Mann-Whitney U test was used to measure statistical differences between the groups. No significant differences were recorded, n=22 for cases and n=20 for controls.

Total GM-CSF cytokine responses were similar between TB-exposed and non-exposed infants: p=0.813, as shown in Figure 1B. The median and IQR are shown in Table 2. We further observed a slight increase in the total response of IFN γ among the TB-exposed versus the TB-non-exposed infants. However, this was not statistically supported: p=0.504, as shown in Figure 1C. The median and IQR are shown in Table 2. This trend was also seen with TNF-α and IL-2 cytokine responses: p=0.623/0.520, as shown in Figure 1D and E, respectively. The median and IQR are shown in Table 2.

#### TB1 cytokine responses

TB1 responses for IL-17A, GM-CSF, IFN-γ, TNF-α, and IL-2 were similar between TB-exposed and non-exposed infants: p=0.150/0.195/0.613/0.696/0.175, as shown in Figures 2A, B, C, D, E, and E, respectively. The median and IQR values are shown in Table 2.

**Figure 2:**
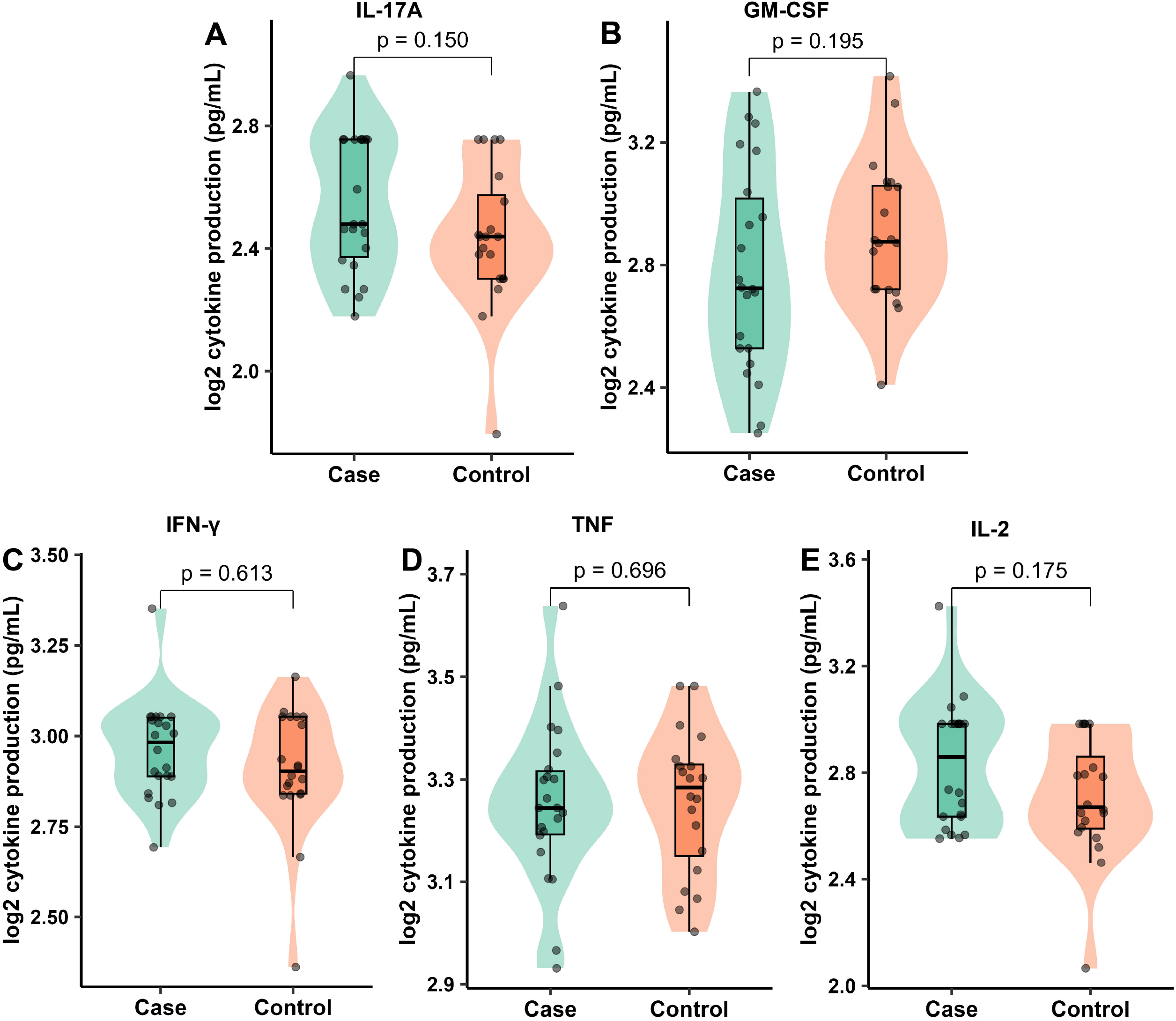
TB1 cytokine responses for IL-17A, GM-CSF, IFN γ, TNF-α, and IL-2 among TB-exposed and TB-non-exposed infants were measured by Luminex. TB1 responses for IL-17A(A), GM-CSF(B), IFN-γ(C), TNF-α(D), and IL-2(E). Box plots show data represented as median and interquartile ranges. Mann-Whitney U test was used to measure statistical differences between the groups. No significant differences were recorded, n=22 for cases and n=20 for controls.

#### TB2 cytokine responses

TB-exposed infants expressed higher levels of IL-17A cytokine responses than non-exposed infants: p=0.049, as shown in Figure 3A. The median and IQR are shown in Table 2. We observed similar responses between TB-exposed and non-exposed infants for cytokines: GM-CSF, IFN-γ, TNF-α, and IL-2: P=0.940/0.623/0.489/0.649, as shown in Figure 3B, C, D and E, respectively. The median and IQR values are shown in Table 2.

**Figure 3:**
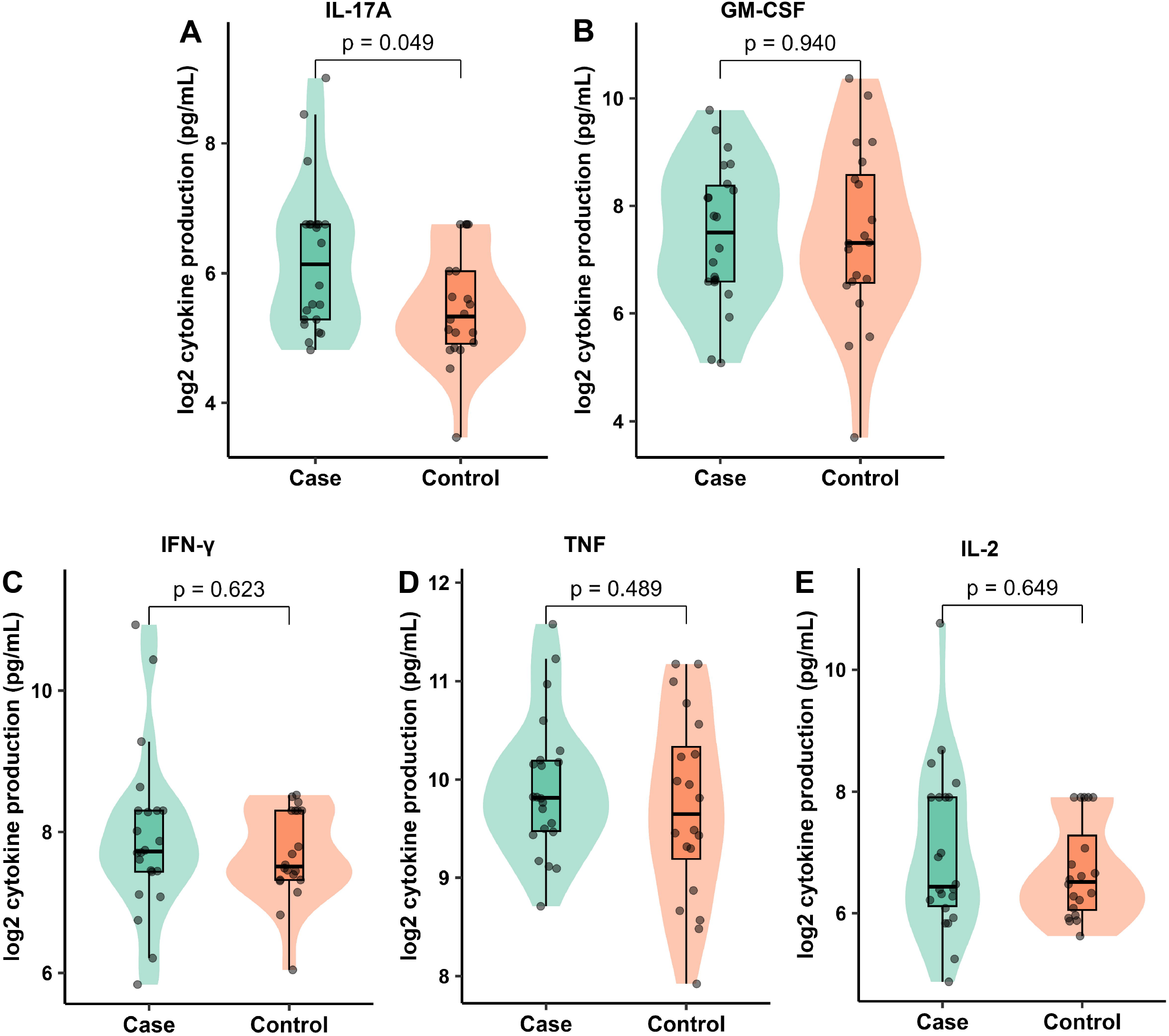
TB2 cytokine responses for IL-17A, GM-CSF, IFN γ, TNF-α, and IL-2 among TB-exposed and TB-non-exposed infants were measured by Luminex. TB2 responses for IL-17A(A), GM-CSF(B), IFN-γ(C), TNF-α(D), and IL-2(E). Box plots show data represented as median and interquartile ranges. Mann-Whitney U test was used to measure statistical differences between the groups. No significant differences were recorded, n=22 for cases and n=20 for controls.

#### Correlation between cytokine responses and haemoglobin concentrations

We measured the correlation between cytokine responses and haemoglobin concentrations between TB-exposed and non-exposed infants. Figure 4A shows the correlation between total cytokine responses and haemoglobin concentrations. We observed a strong positive correlation between TNF-α responses and haemoglobin, implying that an increase in haemoglobin concentration among infants caused a corresponding increase in TNF-α cytokine responses for both TB-exposed and non-exposed infants: p=0.038, R=0.456. Haemoglobin concentrations were not observed to influence responses of TB-exposed and non-exposed infants for the cytokines; however, the correlation was positive: GM-CSF, IL-2, and IFN-γ. GM-CSF had a strong positive correlation, whereas IL-2 and IFN-γ showed weak correlations with haemoglobin concentrations. The P and R values are p=0.092 and R=0.377; p=0.839 and R=0.047; p=0.762 and R=0.07, respectively. IL-17A, on the other hand, showed a negative correlation: p=0.875 and R=-0.037, as shown in Figure 4A.

**Figure 4:**
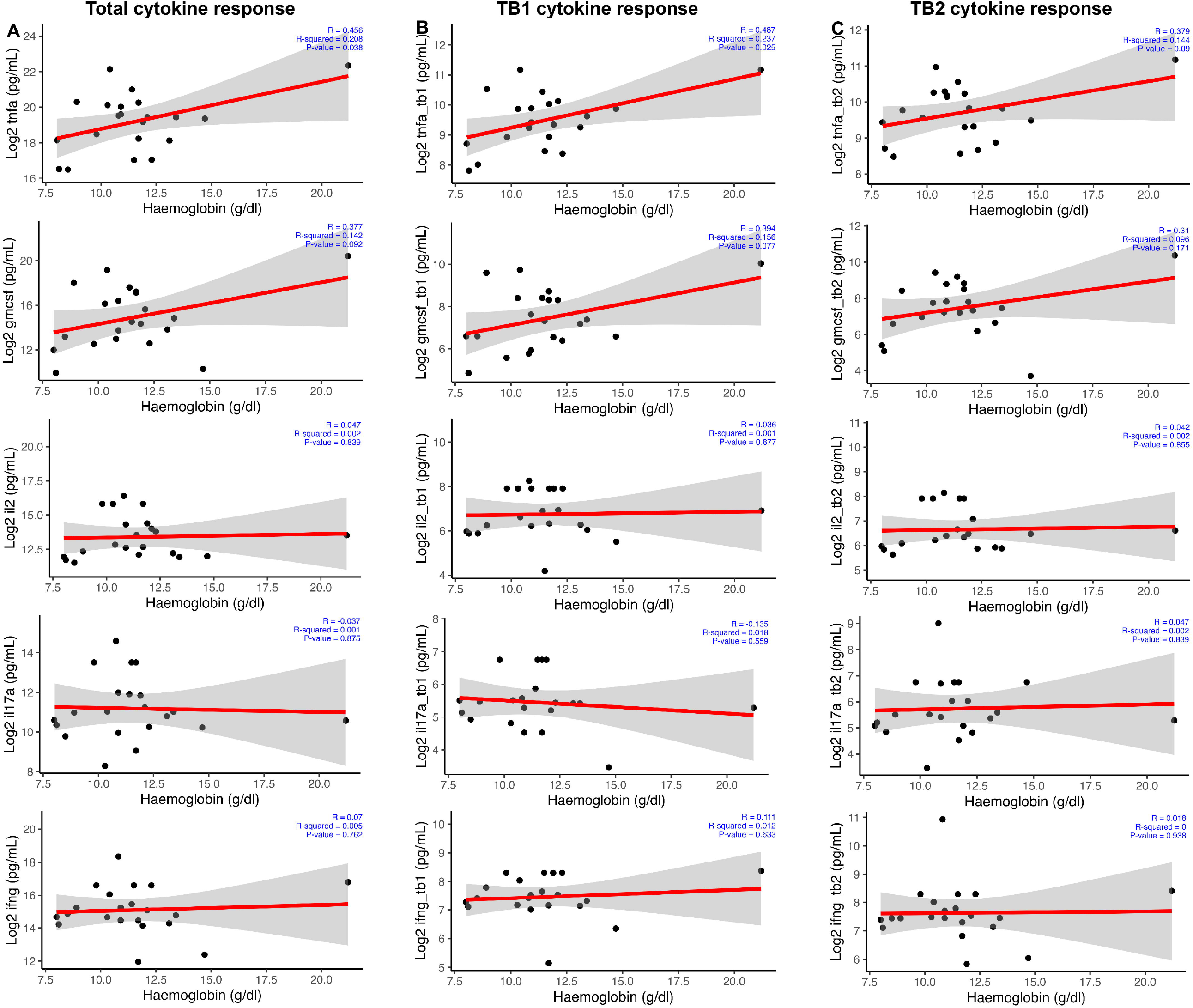
Correlation analysis between cytokine responses and haemoglobin concentrations of infants. Total cytokine response (A), TB1 cytokine response (B), and TB2 cytokine response (C). Scatter plots show data represented as median and interquartile ranges. Mann-Whitney U test was used to measure statistical differences between the groups. No significant differences were recorded, n=22 for cases and n=20 for controls.

Next, we found that the TNF-α correlations versus haemoglobin were persistent in TB1 responses: p=0.025 and R=0.487, as shown in Figure 4B. Despite the observed positive correlation, the observed correlations for GM-CSF, IL-2, and IFN-γ were not statistically significant. GM-CSF had a strong positive correlation, whereas IL-2 and IFN-γ showed weak correlations with haemoglobin concentrations. The P and R values are p=0.077 and R=0.394; p=0.877 and R=0.036; p=0.633 and R=0.111, respectively. IL-17A, on the other hand, showed a negative correlation: p=0.559 and R=-0.135, as shown in Figure 4B.

Finally, we observed positive correlations for TNF-α, GM-CSF, IL-2, IL-17A, and IFN-γ in the TB2 responses. The Pand R values are p=0.09 and R=0.379; p=0.171 and R=0.31; p=0.855 and R=0.042; p=0.839 and R=0.047; p=0.938 and R=0.018 as shown in Figure 4C, respectively.

## Discussion

The immune system mainly relies on cell-mediated immunity to fight against TB. T cells are recognised as the most important in fighting this intracellular pathogen due to the production of inflammatory cytokines like IFN-γ [17]. During active TB infection, there is an additional heightened immune response of proinflammatory cytokines, such as IL-17, TNF-α, and IL-2. When a pregnant woman has active TB, these proinflammatory cytokines may cross the placenta and affect the developing fetus. For example, IL-17 and IFN γ have been implicated in deterring brain development of the fetus in mouse models [18].

In this study, we found that infants exposed to TB had low birth weights compared to TB-non-exposed infants. We also found that TB-exposed infants had low haemoglobin (Hb) concentrations compared to infants not exposed to TB. We suggest this could be due to TB. Moreover, correlations performed for cytokine production versus Hb concentrations for TB-exposed and TB-no-exposed infants in Figure 4 revealed a strong positive correlation between TNF and Hb concentrations.

We found that the TB-exposed infants had slightly increased IFN-γ production compared to the TB-non-exposed infants. Several factors may contribute to this insignificant IFN-γ production. First, maternal immune components may cross the placenta and suppress the immune response of the fetus. For example, regulatory T cells and cytokines such as IL-10, which can inhibit IFN-γ production, may be transferred from the mother to the fetus during pregnancy [18] & [19]. However, we did not analyze the regulatory responses in this study to confirm this anticipation. Evidence suggests that maternal antibodies against *Mtb*, like IgG1, may be transferred to the fetus and interfere with the T cell response to *Mtb* antigens [20] & [21]. Also, the immature immune system of the neonate may be less able to mount an effective immune response to *Mtb* antigens [22]. As a result, the neonatal immune response to *Mtb* antigens may be weaker, and IFN-γ production may be reduced [23]. Additionally, exposure to *Mtb* antigens during fetal development may induce immune tolerance, leading to a reduced immune response to *Mtb* antigens after birth [20]. This is thought to occur by activating regulatory T cells that suppress the immune response to *Mtb* antigens. As a result, the neonatal immune response to *Mtb* antigens may be weaker, and IFN-γ production may be reduced. This reduced IFN-γ production in infants born to active TB mothers may have implications for the development of future risk of TB infection acquisition and progression to TB disease.

The elevated IL-17A cytokine response observed in the TB-exposed infants was of special interest, especially in the TB2 tube, demonstrating CD4 and CD8 T cell responses and possible ongoing or recent *Mtb* presence. This result was also observed in the total responses of both TB1 and TB2 combined. Some studies have associated high IL-17 with the progression of the disease [24], [25], [26] & [27]. We show that these infants have an intensified immune response against TB [28] and may have an increased risk of TB infection or disease progression. These findings may have implications for early diagnosis and intervention strategies in this vulnerable population. Monitoring IL-17 responses could also serve as a potential biomarker for identifying infants at higher risk of TB infection, enabling prompt detection and timely intervention to prevent disease development.

It is worth noting that the TB-non-exposed infants had higher GM-CSF total responses as compared to the TB-exposed infants. Production of GM-CSF by activated T cells is induced by proinflammatory cytokines [29] such as TNF α. The role of GM-CSF in *Mtb* infection is reported to be principally activating macrophages to produce more inflammatory cytokines to induce the differentiation and activation of T cells into Th 17 [14] thereby maintaining a proinflammatory state during infection [30]. One study found that GM-CSF and TNF-α are among the most commonly detected cytokines in the cord blood of healthy new-borns, along with interleukin-1 beta (IL-1β) and interleukin-6 (IL-6) [31] and this is similar to our observation. In newborns, GM-CSF may be necessary for developing the immune system and establishing protective immunity against infections [32] therefore, our results show that the TB-non-exposed infants were healthy.

In this study, we could not assess the anti-inflammatory T-cell responses, which could have impacted our findings. However, we demonstrated significant trends that can be utilised for additional studies.

In conclusion, this study provides evidence that infants born to active TB mothers exhibit heightened Th 1 T cell responses and more markedly IL-17 responses compared to infants born to mothers without active TB. These findings shed light on the immune responses in this specific vulnerable population. They may aid in the development of improved strategies for the early detection and management of TB in newborns at higher risk.

## Data Availability

Raw data that guided these conclusions will be made available by the authors upon request.

## Abbreviations

BCG: *Bacille Calmette-Guérin*)
DPT: Diphtheria-Pertussis-Tetanus)
IFN-γ: InterFeron Gamma)
Mtb: Mycobacterium tuberculosis)
IL-2: Interleukin 2)
TNF-α: Tumour Necrosis Factor Alpha)
IL: 17A-Interleukin 17A)
GM: CSF-Granulocyte Macrophage-Colony Stimulating Factor)
QFT: QuantiFERON)
TB: Tuberculosis)
ESAT-6: Early Secreted Antigenic Target 6)
CFP-10-: Culture Filtrate Protein 10)
IQR: Interquartile range)

## Declarations

### Ethical consideration

This study received ethical approval from the School of Medicine Research and Ethics Committee (SOMREC reference number SOM 2020-11); the School of Biomedical Sciences (SOBSREC reference number SBS-2022-226) at Makerere University; and the Uganda National Council for Science and Technology (UNCST registration number HS1396ES). The mothers provided written informed consent and assent for their babies to participate in the study.

### Data availability

Raw data that guided these conclusions will be made available by the authors upon request.

### Author contributions

DS performed laboratory experiments, analyzed data, and drafted the manuscript. IAB, AN, SC, RN, FB, JBB, PS, APK, DK, and NR participated in the concept development and reviewed the manuscript. PS developed the experimental protocols that followed the experiment and offered technical laboratory training to perform the experiments. IAB sourced funding for the research.

## Acknowledgment

A vote of special thanks goes to the Tuberculosis and Comorbidities Research Consortium’s field team for data collection. We thank the study participants and the health facility administrators for their engagement in this study. This work was supported by the Crick African Network, which receives its funding from the UK’s Global Challenges Research Fund (MR/P028071/1), and by the Francis Crick Institute, which receives its core funding from Cancer Research UK (FC1001647), the UK Medical Research Council (FC1001647), and the Wellcome Trust (FC1001647). We also acknowledge HEPI-SHSS for offering a research contribution to the laboratory work.

